# Controlling SARS-CoV-2 in schools using repetitive testing strategies

**DOI:** 10.1101/2021.11.15.21266187

**Authors:** Andrea Torneri, Lander Willem, Vittoria Colizza, Cécile Kremer, Christelle Meuris, Gilles Darcis, Niel Hens, Pieter Libin

## Abstract

SARS-CoV-2 remains a worldwide emergency. While vaccines have been approved and are widely administered, these are only available to adults and adolescents in Europe. Therefore, in order to mitigate the spread of more transmissible SARS-CoV-2 variants among children, the use of non-pharmaceutical interventions is still warranted. We investigate the impact of different testing strategies on the SARS-CoV-2 infection dynamics in a primary school environment, using an individual-based modelling approach. Specifically, we consider three testing strategies: 1) *symptomatic isolation*, where we test symptomatic individuals and isolate them when they test positive, 2) *reactive screening*, where a class is screened once one symptomatic individual was identified, and 3) *repetitive screening*, where the school in its entirety is screened on regular time intervals. Through this analysis, we demonstrate that repetitive testing strategies can significantly reduce the attack rate in schools, contrary to a reactive screening approach. Furthermore, we investigate the impact of these testing strategies on the average number of school days lost per child.

## 1 Introduction

The SARS-CoV-2 pandemic has caused over 200 million COVID-19 cases and over 4 million deaths around the world up to September 2021 [23]. Although vaccines have been approved and are widely administered, to date, these vaccines only received approval for its use in adults and adolescents in Europe (i.e., 12+ years old) [9]. While the contribution of children in the COVID-19 epidemic is still subject to ongoing debate [10], there is a consensus that more infectious variants can cause significant outbreaks among children [2]. Furthermore, recent work indicates that children, who typically undergo an infection with little or no symptoms, might still be highly contagious and as such generate new infections in the community [17]. In the absence of a licensed vaccine for children, the only means to mitigate outbreaks of SARS-CoV-2 in primary schools, is through non-pharmaceutical interventions, including the use of masks, social distancing, hygienic precautions and diagnostic testing. Here, the aim of diagnostic testing is to detect and subsequently isolate infected individuals. Therefore, it is important to advance our understanding on how different testing strategies impact primary schools, considering the evolution of SARS-CoV-2 contagiousness through different variants of concerns (VoCs). When defining intervention policies in a school setting, attention needs to be devoted to limiting the number of school days lost. In fact, Engzell et al. evaluated the impact of school closures on students’ learning performance, finding that students made less progress while learning from home [8]. In this work, we use a model-based approach to evaluate different testing strategies in a primary school context, to assess the impact of such strategies on the school environment in terms of disease spread and the average number of school days missed per child. To this end we construct an individual-based model that explicitly represents a set of primary school pupils. These pupils are allocated to a fixed set of classes and are taught by a fixed set of teachers. Through this micro-model, we perform a fine-grained evaluation of testing strategies, keeping track of both the attack rate and the number of school days lost. We conduct experiments considering different *R*_0_ values to reflect the increase in infectiousness exhibited by the most recent Delta VoC.

Through our modelling experiments we present two major findings. Firstly, we demonstrate that repetitive testing policies can significantly reduce the attack rate in schools. Secondly, we find that a purely reactive testing approach, where a class is only screened once a symptomatic individual was identified, is an ineffective policy to reduce the attack rate given the asymptomatic nature of SARS-CoV-2 infections in children, and such strategies might confer a false sense of security. Simulation results are consistent throughout the different simulated settings, in which the assumptions on the model parameter values are challenged in a sensitivity analysis. Furthermore, our model and experimental results provide a substantial basis for subsequent analysis on how to react to future pandemics in schools, as we show the impact of the proportion of asymptomatic infection on testing strategies.

## 2 Individual-based primary school model

We construct an individual-based model to describe COVID-19 outbreaks in a primary school setting, which we briefly introduce in this section ^1^. Children are assigned to classes and we simulate interactions among children both within and between classes. Teachers are allocated to specific classes and are assumed to interact only with individuals with whom they share the same class. We assume that symptomatic individuals develop symptoms at the peak of their infectiousness, at which they can be detected and placed in isolation for 10 days. We implement three testing policies aimed at mitigating school outbreaks:

- *Symptomatic Isolation* (SI). Symptomatic individuals are detected with probability *p*_*D*_ and tested. Individuals that test positive are put in isolation.
- *Reactive Screening* (ReaS). Symptomatic individuals are detected with probability *p*_*D*_ and tested. Individuals that test positive are put in isolation. In addition, all members of the class where this case originates from are also tested. When any additional cases are detected, these individuals are also put in isolation.
- *Repetitive Screening* (RepS). All of the school’s members are tested on a repetitive basis. All individuals that test positive are put in isolation.

All testing policies will close a class when the number of infections in this class exceeds 2 cases. Analogously, all testing policies will close the school when the number of infections over all classes exceeds 20 cases. Infection counts are recorded in a 14 day time window to determine class and school closures. These assumptions are challenged in a sensitivity analysis, which we discuss in the Results section.

### Experimental framework

Model parameters are set to describe COVID-19 spreading. We represent both the Wuhan strain of SARS-CoV-2 and the Delta variant by setting different transmission potentials given a contact, informing such values from the literature [13, 4]. We consider a distinct detection probability of symptoms *p*_*D*_ for children (*p*_*D*_ = 0.3) and adults (*p*_*D*_ = 0.5), as children typically exhibit such mild symptoms that are easily overlooked [18]. Children are set to be halve as susceptible as adults [7]. We assume that 30% of school children are immune due to prior infection ^2^, and that 90% of the teachers are immune, due to their vaccination status or due to prior infection.

The simulated testing procedure accounts for the use of PCR tests on saliva or throat washing samples. The sensitivity of such tests is set to 86%, and there is a one day delay in reporting the result [5]. Recent reports show that the performance of saliva sampling in combination with PCR testing is on par with nasopharyngeal swab sampling in combination with PCR testing [24]. We assume full compliance to testing, that could potentially be reached since saliva sampling is less invasive compared to other specimen collection procedures. Infectious individuals become PCR detectable 2 days after infection, as previously assumed [20, 21]. For the reactive screening testing policy, we assume that there is a one day screening delay.

Every simulated week, five susceptible children are assumed to acquire infection outside the school environment, accounting for disease importation or seeding. The epidemic is simulated for 100 days and we consider 100 simulation runs to present our final results. In each simulated outbreak, we keep track of the total number of infections and the total number of school days lost.

## 3 Results

To investigate the efficacy of the testing strategies, we consider both the attack rate^3^ and the average number of school days lost per child (NSDL). In order to differentiate between the initial and current phase of the epidemic, we compare two scenarios that represent the Wuhan strain and the Delta VoC, characterized by a different transmission potential given a contact. While the overall trends of the reported measures are similar for the Wuhan and Delta scenarios, the difference between the testing strategies is most pronounced in case of a more infectious virus strain (i.e., the Delta VoC).

On the one hand, our experiments show (Figure 1) that symptomatic isolation results in the infection of a significant proportion of the school population. This comes as no surprise, as in our model we assume that 80% of the pupils will go through the infection asymptomatically, and by following this testing policy, we are only able to pick up infections that make up the tip of the iceberg. On the other hand, the attack rate consistently decreases when a testing policy is used that performs a wider screening of the school population, such as reactive testing and repetitive testing. Such policies enable the detection of both symptomatic and asymptomatic cases, that can be subsequently isolated, thereby limiting further transmissions. However, contrary to intuition, our experiments indicate that the reactive screening strategy performs only slightly better than symptomatic isolation. This can be explained by the low probability that pupils will be symptomatic when infected, hence a low probability to trigger the reactive screening. When we assume that 80% of infections in children progress asymptomatically, we can expect that four asymptomatic generations take place, on average, before a symptomatic infection is observed^4^. Therefore, when a reactive screening procedure is triggered by a symptomatic individual, the infected individuals that share a class with this individual might already be recovered or in the end phase of their infectious period. Hence, on average, only a few generations can be avoided by employing a reactive screening strategy, when the infection is predominantly driven by asymptomatic carriers. Note that we also assume that only a limited percentage of symptomatic children is detected (30%), due to the fact that many children exhibit only minor symptoms.

**Figure 1:**
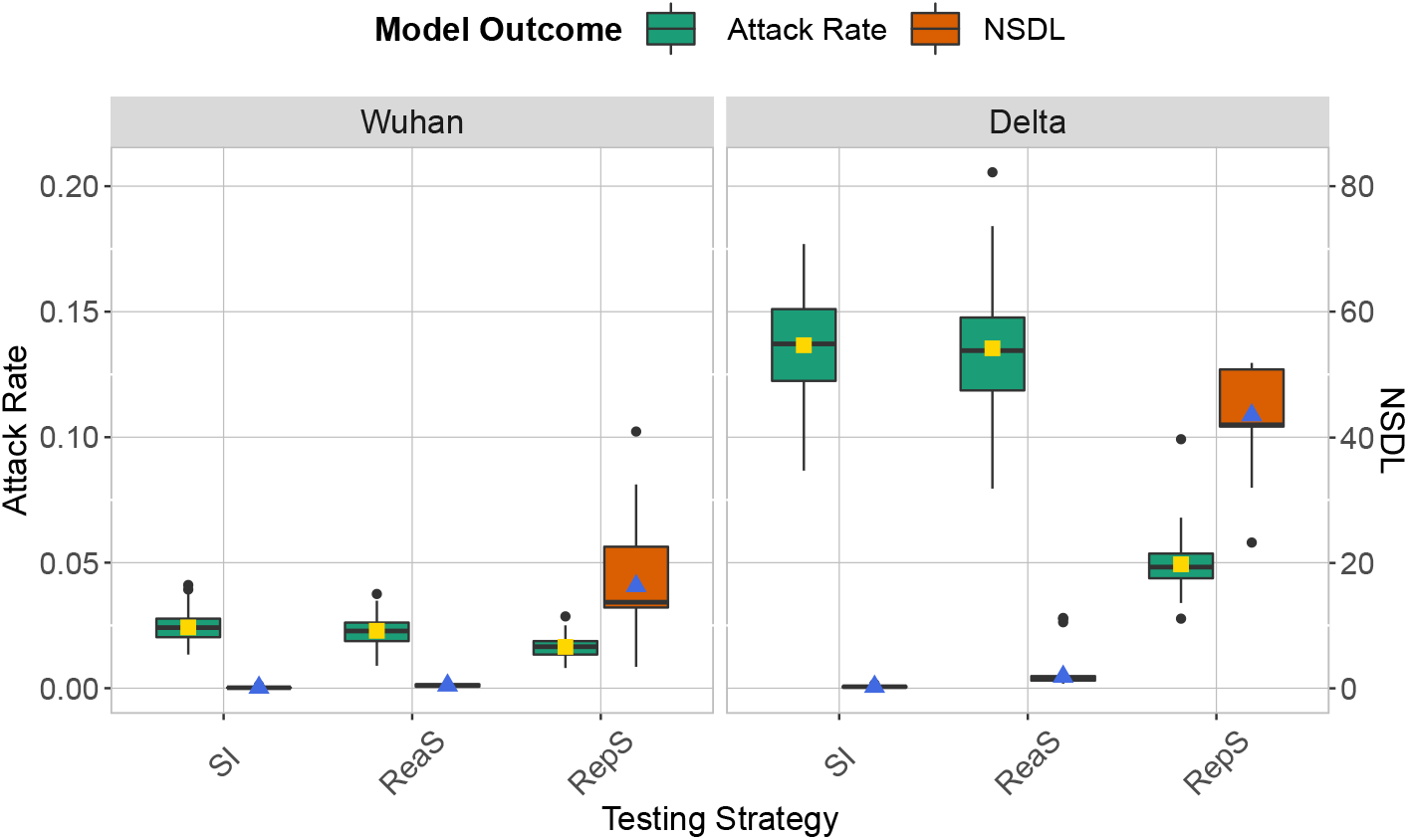
We show the base scenario for the Wuhan strain (left panel) and Delta VoC (right panel) for a moderate seeding of 5 seeds per week. In each panel we consider three testing strategies: symptomatic testing (SI), symptomatic testing in combination with reactive screening (ReaS) and repetitive screening (RepS). For each of the testing strategies we show a boxplot of the attack rate (green boxplot) and NSDL (orange boxplot) together with their mean values (respectively, yellow and blue dots).

To interpret the experimental results with respect to the average number of school days lost per child (NSDL), we need to recognize that children can miss school due to isolation when infected or due to quarantine due to a high risk contact. For the symptomatic isolation strategy, only children with symptoms are isolated, resulting in an average NSDL per child that is directly proportional to the fraction symptomatic cases. For reactive screening, additional asymptomatic pupils might be identified, thereby quickly reaching the class or school closure thresholds, with a higher NSDL as a result. This effect is most pronounced when we apply repetitive testing, where we effectively detect a high proportion of the infections, thereby rapidly meeting the class and/or school closure thresholds, with a very high NSDL as a consequence.

We note that the high NSDL associated with repetitive testing, renders this testing policy impractical. However, we observe that the class and school closure thresholds were selected in order to avoid major outbreaks. We argue that, by using repetitive testing, more lenient thresholds could be applied, as we are able to detect a larger proportion of cases. We investigate this in Figure 2 where we remove the school closure threshold, and investigate a set of class closure thresholds. This experiment confirms that a larger threshold can be used, with only a limited impact on the attack rate, and that such thresholds result in a more acceptable NSDL.

**Figure 2:**
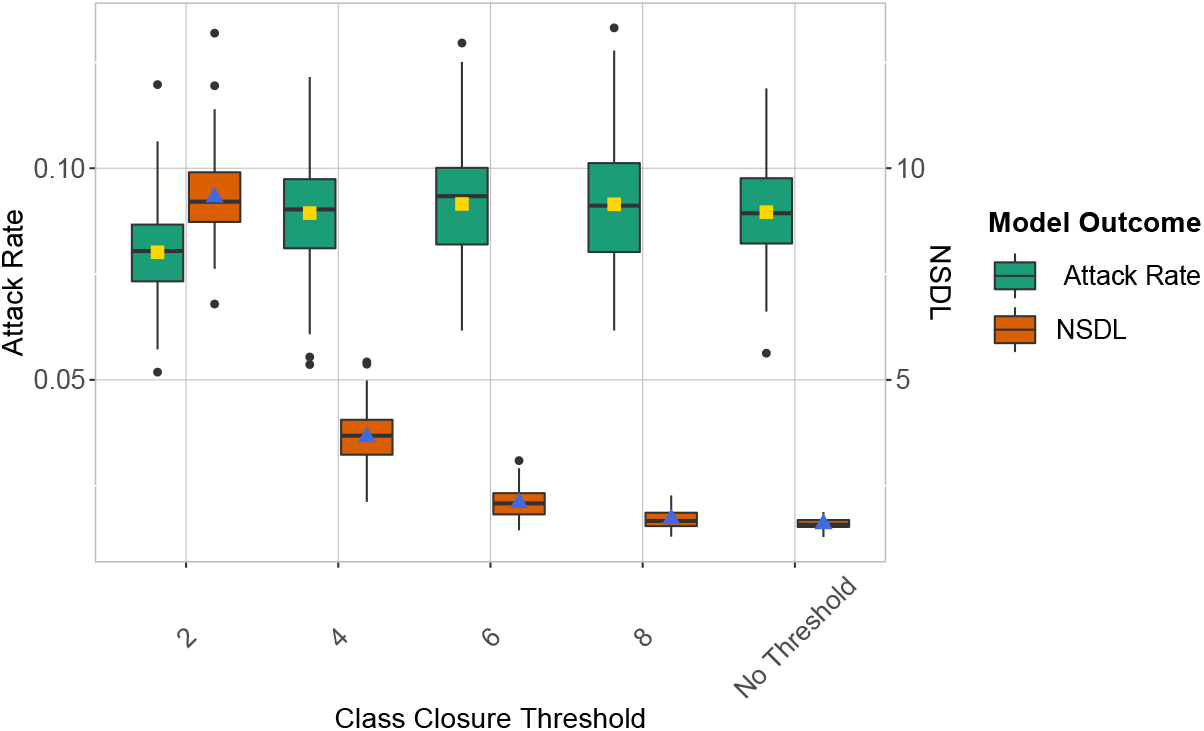
We show the repetitive testing strategy in the context of the Delta VoC for a moderate seeding of 5 seeds per week, where we consider different class closure thresholds, and no school closure threshold. This experiment shows that a higher class closure threshold has little effect on the attack rate, yet it significantly reduces the NSDL.

In order to challenge some of the assumptions of this study, we conduct a series of sensitivity analyses. We show these results in the Supplementary Information and briefly report the main findings. We investigate the impact of the amount of weekly introductions, by seeding 1 and 10 cases on a weekly basis, next to the baseline scenario of 5 cases. We note that the impact of additional seeding cases amplifies the attack rate, but overall repetitive testing proved to be robust in regard of this parameter. Furthermore, the high efficacy of repetitive testing is also observed when varying the level of contact reduction between classes, and when considering different levels of immunity in children and adults. Next, we assume that asymptomatic individuals are as infectious as symptomatic individuals, as recently observed by Meuris et al. [17]. Also in this case the trends of attack rate and number of school days lost among the different testing strategies are consistent with the baseline scenarios reported above. Finally, we consider a repetitive testing scenario where we test the entire school population twice per week, which shows that such a strategy squashes even an highly contagious epidemic (Figure 3).

**Figure 3:**
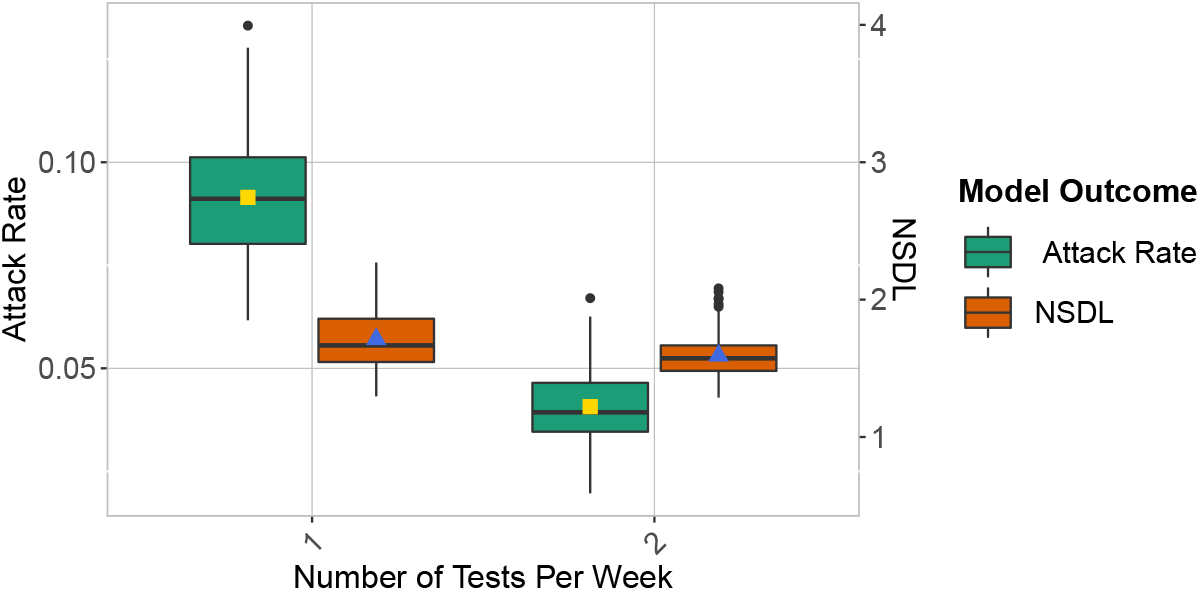
We show the repetitive testing strategy in the context of the Delta VoC for a moderate seeding of 5 seeds per week, where we consider a repetitive testing strategy where the entire school population is tested either once or twice per week. We consider class closure threshold of 8 and no school closure threshold. This experiment demonstrates that even for a highly infectious virus strain, repetitive testing can almost nullify the number of transmissions at school.

## 4 Discussion

This simulation study compares the efficacy of testing strategies for mitigating COVID-19 outbreaks in a school setting. Throughout all simulated scenarios, a repetitive testing procedure is shown to be most efficient to reduce the attack rate. Simulations indicate that such a testing strategy limits the number of transmission events even when no class and school closures are in place. The low efficacy of the symptomatic testing and reactive screening procedures is explained by the asymptomatic nature of SARS-CoV-2 infections, especially for children. In fact, when surveillance is based just on the onset of symptoms, asymptomatic carriers avoid detection and intervention, sustaining the spread of the virus.

Class and school closures affect the number of school days lost of healthy children. To limit the learning loss caused by such closures, a control strategy in which only infected cases are isolated would be optimal. With our experiments, we show that a repetitive testing strategy can achieve this. While keeping transmissions under control, a limited number of school days lost is computed when no thresholds are in place.

In our experiments, we consider PCR tests as golden standard, as we argue that the available testing infrastructure is most appropriate for performing reactive and repetitive screening procedures [14]. To make this procedure more efficient, a class pooling approach could be used to reduce the number of samples to be analyzed [14]. However, we argue that in order to implement pool-based testing on a large scale, a population-based evaluation of the sensitivity of pooled PCR testing is warranted [14]. To further reduce the number of required PCR tests, the use of a repetitive testing strategy can be targeted to areas where prevalence is particularly high.

The presented simulation model was set to describe the spreading of COVID-19. However, other infectious diseases can easily be represented, incorporating the specific transmission characteristics of the pathogens in the simulator. Especially in the case of emerging epidemics or pandemics with higher contagiousness in child-to-child interactions and/or a higher severity for children, an appropriate testing strategy in a school setting is pivotal to dampen epidemic spread. By using the developed model, ad-hoc testing strategies can be simulated offering valuable insights in controlling epidemics. Furthermore, an additional range of scenarios could be simulated, gathering information on the best policy that needs to be applied in specific epidemiological contexts.

Other work has shown that viral load for symptomatic and asymptomatic carriers is similar [17], indicating that their transmission potential is the same. We tested this assumption noticing an overall increase in the attack rate values. Our experiments confirm that repetitive screening procedures reduce the risks of school transmissions in this scenario.

We assume that teachers are allocated to specific classes and are assumed to interact only with individuals with whom they share the same class. This means that the interaction between teachers in the school environment is limited. We argue that this is a reasonable assumption at this stage of the epidemic, where a large proportion of teachers is either immune or vaccinated. In order to add such functionality to the model, an additional contact structure could be added to the model in which teachers meet, i.e., a teacher room, to be informed by the contact frequencies between adults in a school environment [22].

In this work, we assume perfect compliance by school individuals for both the reactive and repetitive screening. We argue that this is a reasonable assumption, as the threshold for participating in saliva sampling is low, and societal awareness and support for this policies can be achieved, via prompt governmental communication. Nonetheless, we acknowledge that evaluating reduced compliance is interesting to consider, as a means to monitor the epidemic situation when prevalence is low, or to reduce the number of required tests, when such tests are only scarcely available, e.g., at the start of a pandemic. Compliance of testing can be incorporated in the simulator by randomly sampling a proportion of the school population, prior to testing.

## Data Availability

All results presented in this work can be produced with the code on https://github.com/AndreaTorneri/TestingStrategie

## 5 Availability of data and materials

Source code of the individual-based model was implemented in R and is available in the GitHub repository at the following link: https://github.com/AndreaTorneri/TestingStrategies.

## 6 Funding

L.W and P.J.K.L. acknowledge support from the Research Foundation Flanders (FWO, fwo.be) (postdoctoral fellowships 1234620N and 1242021N).

N.H. and P.J.K.L. acknowledge support from from the European Research Council (ERC) under the European Union’s Horizon 2020 research and innovation programme (grant number 101003688—EpiPose project).

N.H. and A.T. received funding from the European Research Council (ERC) under the European Union’s Horizon 2020 research and innovation programme (grant number 682540—TransMID project). N.H. acknowledge funding from the Antwerp Study Centre for Infectious Diseases (ASCID) and the chair in evidence-based vaccinology at the Methusalem-Centre of Excellence consortium VAX-IDEA.

C.M. received funding from the “Fondation Léon Fredericq” and the “Fond d’investissement de recherche scientifique” from the CHU of Liège.

G.D. received “Post-doctorate Clinical Master Specialists” funding from the Fund for Scientific Research (F.R.S.–FNRS, frs-fnrs.be).

We used computational resources and services provided by the Flemish Supercomputer Centre (VSC), funded by the FWO and the Flemish Government.

The funding agencies had no role in study design, data collection and analysis, decision to publish, or preparation of the manuscript.

## 7 Competing Interests

All the authors declare that no competing interests exist.

## Supplementary information

### Individual-based school model

#### Transmission model

We extend the SARS-CoV-2 transmission model presented by Torneri et al. [21] to investigate school settings. In this model, the infection dynamic is described as follows. Individuals are initially susceptible and once infected, they enter the exposed stage. The infection can be asymptomatic or symptomatic. Symptomatic individuals develop symptoms after a pre-symptomatic period. Each symptomatic individual is assumed to show symptoms at the peak of their infectiousness, as indicated by literature findings [19, 1]. After infection, individuals will eventually recover, after which they are assumed to be immune to reinfection.

Infection events are simulated with a counting process approach. First, contacts between individuals are generated. Contacts are effectives, i.e. lead to the transmission of the virus, according to a Bernoulli trial, based on the time since infection. Effective contacts that take place between susceptible and infectious individuals result in infection events. The probability that a contact is effective is composed of two factors: the infectivity measure, *ν*(*t*) and the transmission potential *q*, which accounts for the transmissibility of the pathogen and the susceptibility of the exposed individual. In this context, the basic reproduction number of an infectious disease is approximated with the mean number of effective contacts an infectious individual generates in a fully susceptible population throughout his or her infectious period[21].

The infectivity measure *ν*(*t*) is defined over the exposed and infectious period of the infected individual and is set to represent the shape of the viral load curve for a SARS-CoV-2 infection, under the assumption that a higher amount of virus corresponds to a higher transmission probability [3]. Based on literature findings, we define an infectivity measure that peaks at symptom onset and lasts 10 days, on average [25, 26, 12, 16, 15, 1, 6]. In addition, *ν*(*t*) has an initial plateau with value zero that accounts for the exposed phase.

Asymptomatic and symptomatic individuals are assumed to have the same viral progression, as argued in [27, 26], but we introduce a different level of infectiousness between infectious individuals based on the clinical outcome. Precisely, the relative infectiousness of asymptomatic compared to symptomatic is 0.5 [7].

#### School classes, pupils and teachers

We consider a population of children in primary school (6-12 years old), where each child is randomly allocated to a class. To this end, we construct a set of classes of which the size is sampled from a probability mass function informed by a survey on Belgian primary schools^5^, up until at least 1000 pupils are allocated to these classes.

In the school, we consider teaching and supportive staff, to which we will refer as teachers from this point forward for brevity. The number of 9 teachers is proportional to the number of pupils (ratio 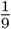)^6^. We consider different contact ratios within *λ*_*w*_ and between *λ*_*b*_ classes, and assume that teachers have contacts with children and other teachers of their allocated classes. The within and between classes contact rates for children are set accordingly to a contact data survey that took place in Belgium [11]. The within class contact rate is given by number of contacts that take place in primary schools and last more than 1 hr (*λ*_*w*_ = 6.62). The between class contact rate is computed as the the number of contacts that last less than 1 hr (*λ*_*b*_ = 2.5). However, we assumed that in a pandemic setting the number of between class contacts is reduced. In the baseline scenario, we assumed that the between contact rate in a COVID-19 pandemic scenario is 30% of *λ*_*b*_. We test such assumption in the sensitivity analysis by varying this proportion among 20%, 50% and 90%.

### Sensitivity analysis

#### Seeding Number

**Figure 4:**
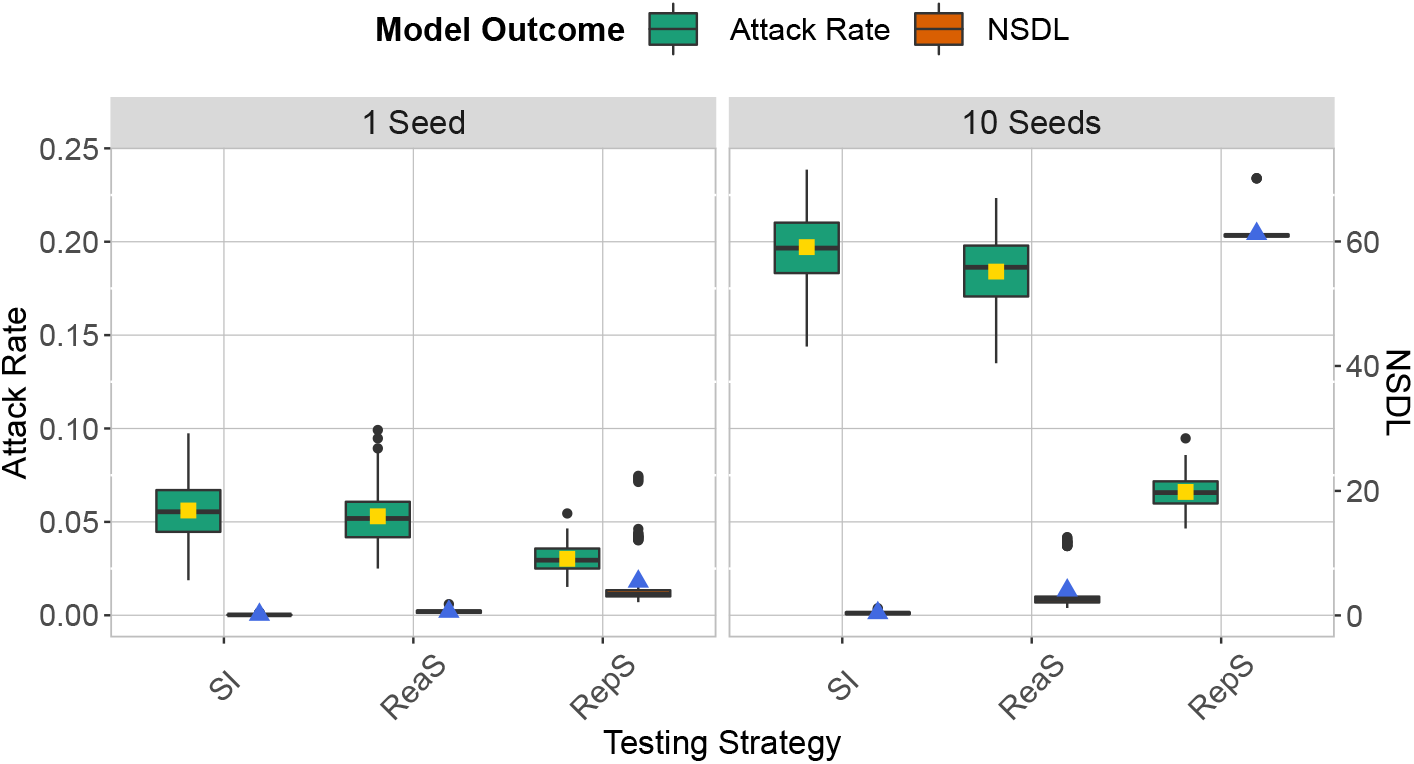
We show the attack rate and the number of school days lost per child when 1 seed (left panels) or 10 seeds (right) are weekly introduced for the Delta VoC. In each panel we consider three testing strategies: symptomatic testing (SI), symptomatic testing in combination with reactive screening (ReaS) and repetitive screening (RepS). For each of the testing strategies we show a boxplot of the attack rate (green boxplot) and NSDL (orange boxplot) together with their mean values (respectively, yellow and blue dots). School and class thresholds are set, respectively, to 20 and 2 detected cases.

#### Number of tests per week

**Figure 5:**
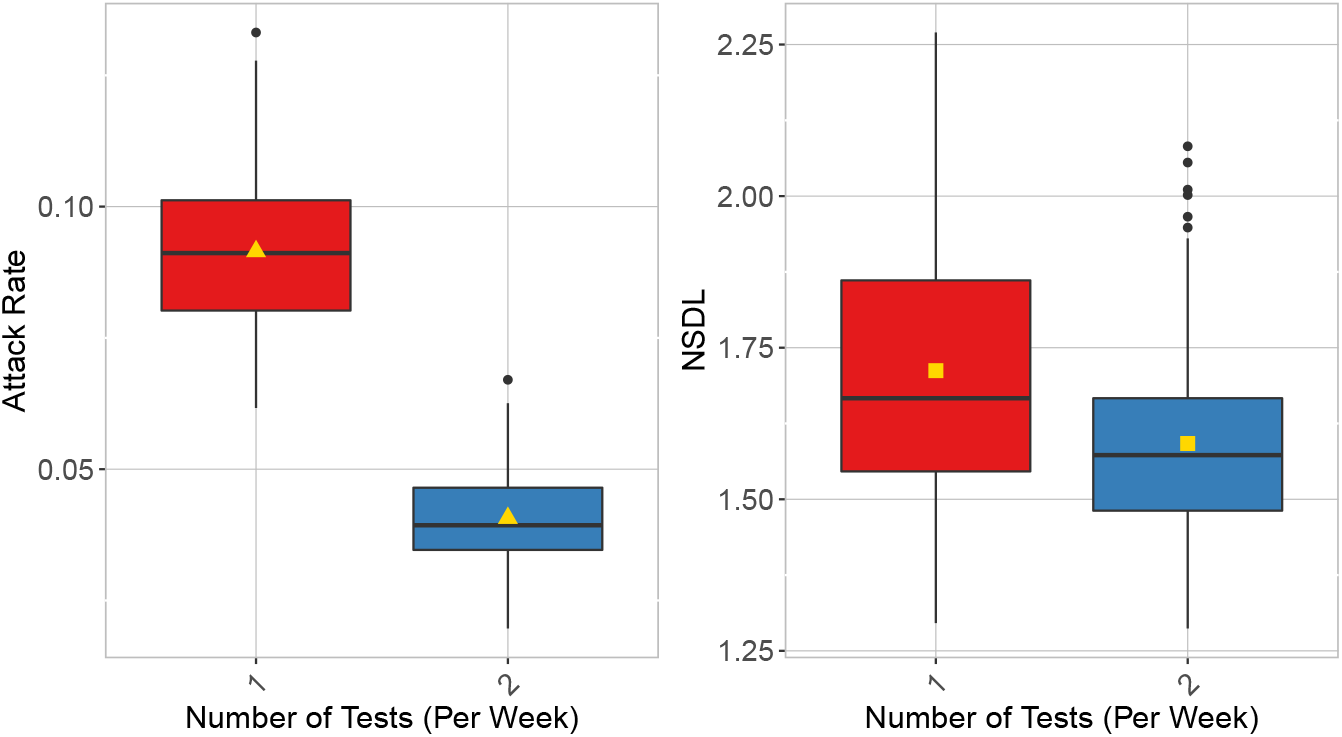
For the repetitive testing strategy, we compare the number of tests used per week in the context of the Delta VoC for a moderate seeding of 5 seeds per week. The class closure threshold is of 8 detected cases, and there is no school closure threshold. This experiments shows that testing twice per week significantly reduces the attack rate (left panel). In addition, the lower number of infections registered, leads to a lower number of NSDL (right panel).

#### Between-classes contact rate

**Figure 6:**
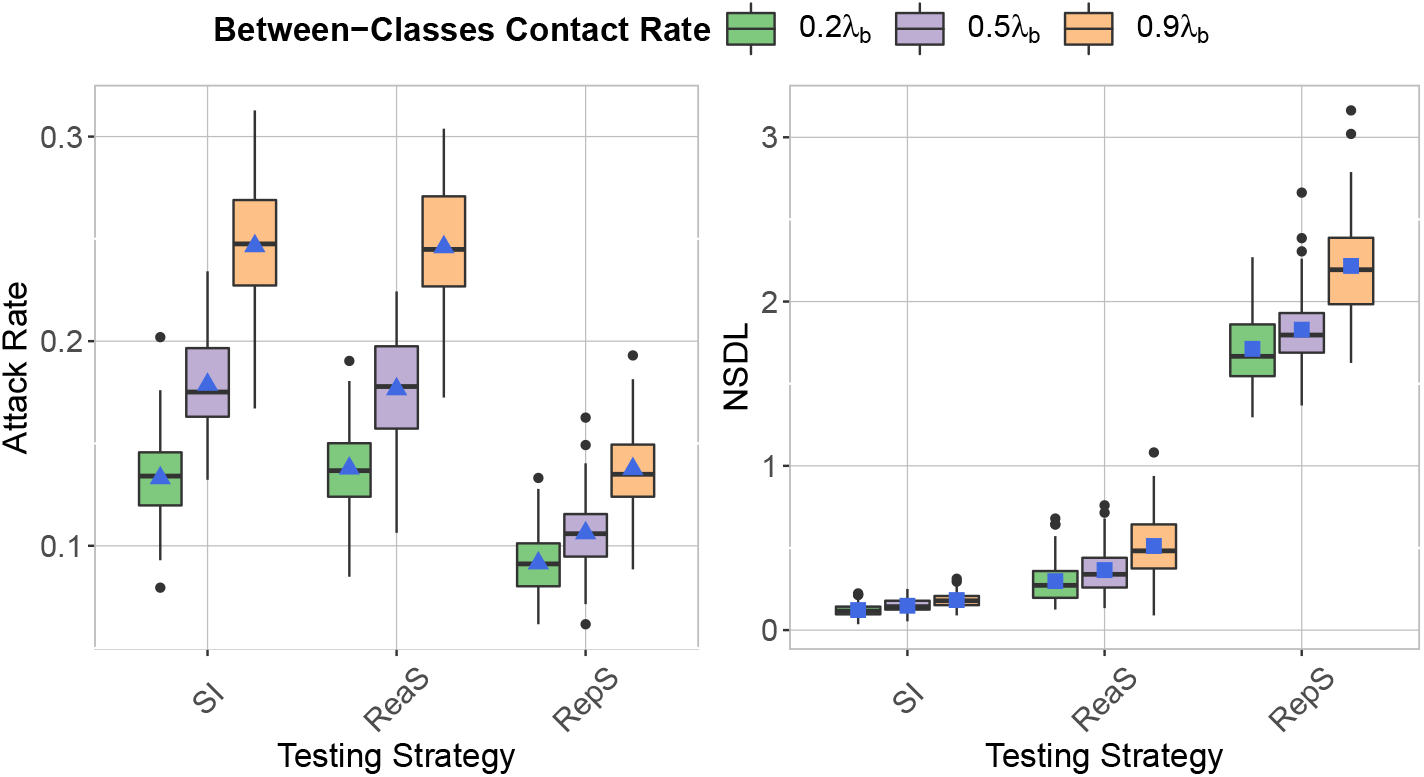
We compare testing strategy in the context of the Delta VoC for a moderate seeding of 5 seeds per week, when varying the proportion of between classes contacts compared to a pre-pandemic scenario. The class closure threshold is of 8 detected cases, and there is no school closure threshold. This experiments shows that an increasing between classes contact rate increases both the attack rate (left panel) and the NSDL (right panel).

#### Immune Population Proportion

**Figure 7:**
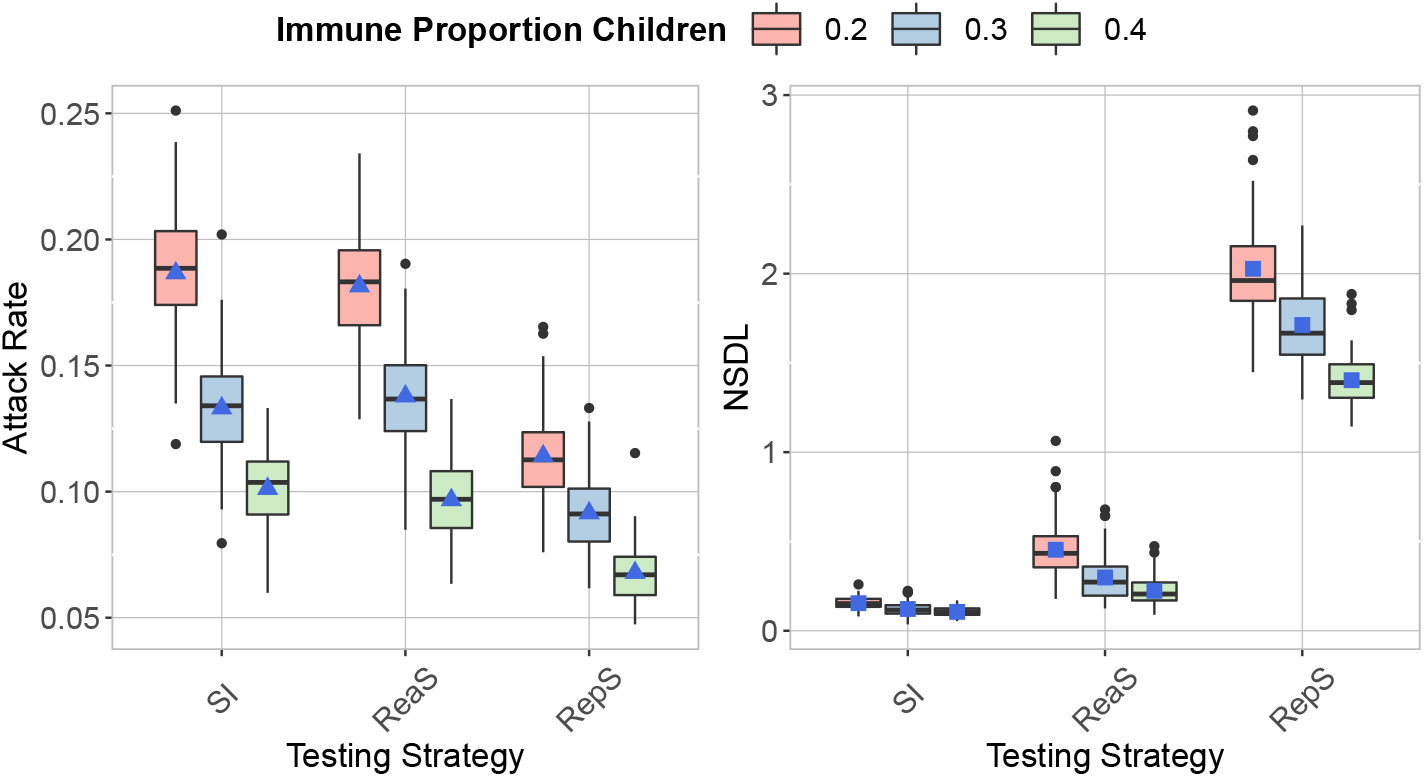
We compare the testing strategies in the context of the Delta VoC for a moderate seeding of 5 seeds per week, when varying the proportion of immune children. The class closure threshold is of 8 detected cases, and there is no school closure threshold. The repetitive screening strategy is shown to decrease the attack rate (left panel) compared to reactive screening and symptomatic isolation, while increasing the NSDL (left panel).

**Figure 8:**
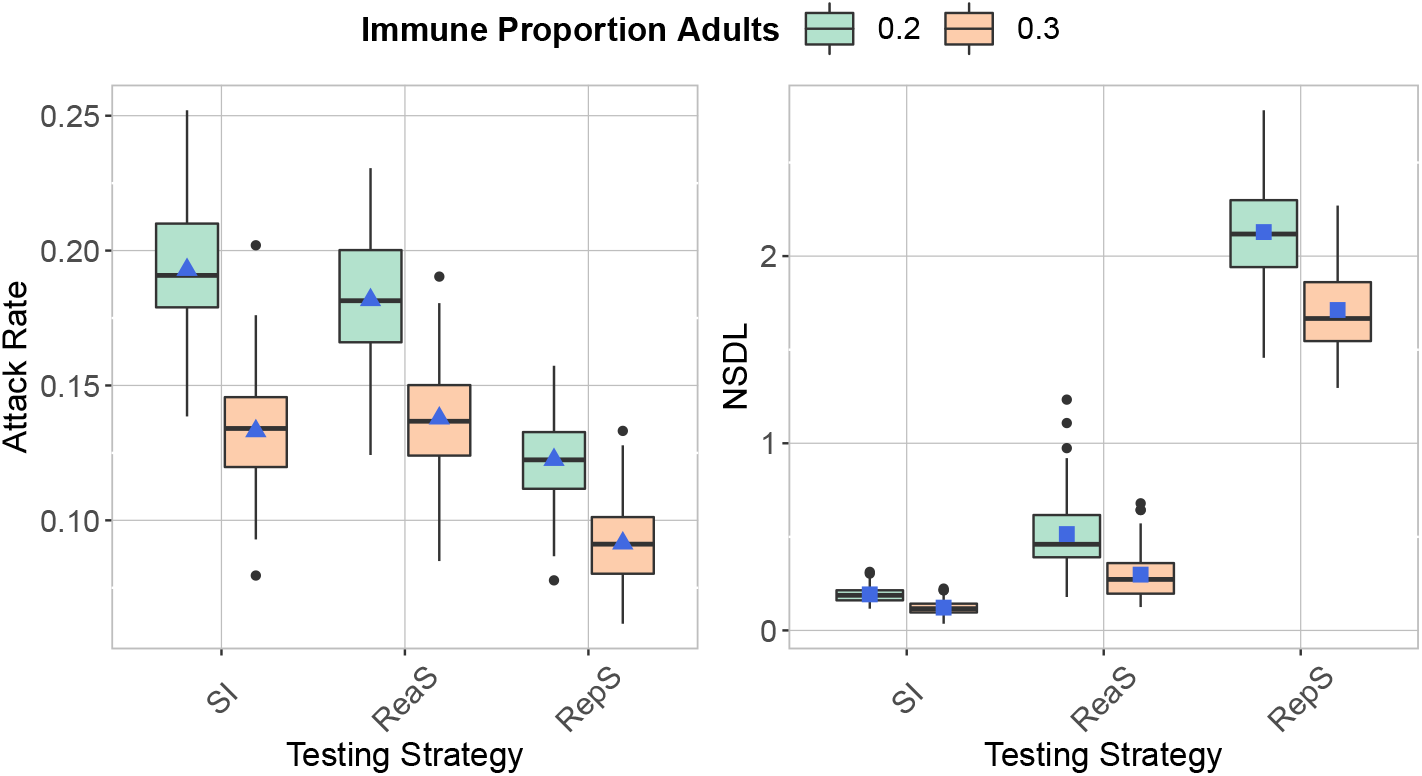
We compare the testing strategies in the context of the Delta VoC for a moderate seeding of 5 seeds per week, when varying the proportion of immune adults. The class closure threshold is of 8 detected cases, and there is no school closure threshold. The repetitive screening strategy is shown to decrease the attack rate (left panel) compared to reactive screening and symptomatic isolation, while increasing the NSDL (left panel).

#### Increased Infectivity Asymptomatic Carriers

**Figure 9:**
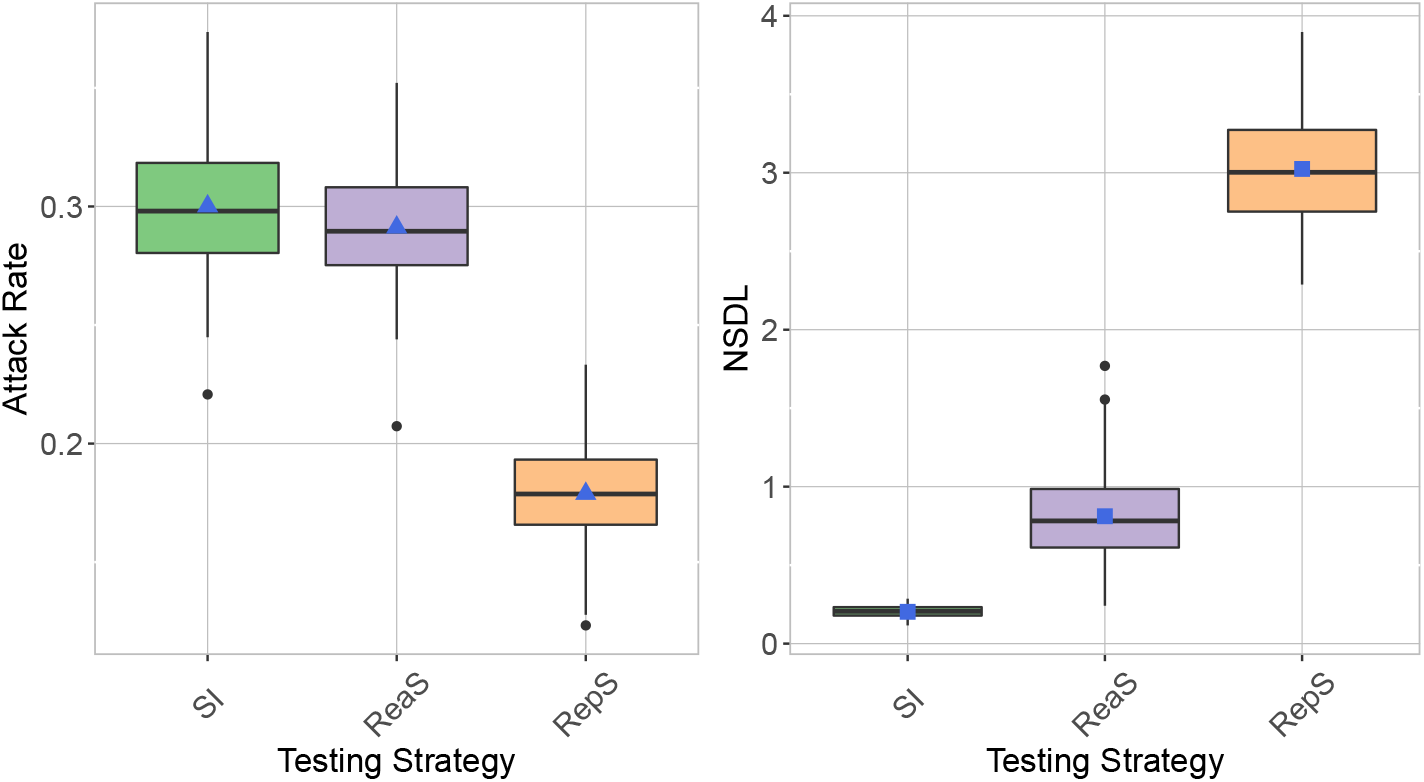
We show the testing strategies in the context of the Delta VoC for a moderate seeding of 5 seeds per week, where we consider asymptomatic carriers to be as infectious as the symptomatic ones. The class closure threshold is of 8 detected cases, and there is no school closure threshold. The repetitive screening strategy is shown to reduce the attack rate (left panel), while leading to a higher NSDL (right panel).

#### School Closure Threshold

**Figure 10:**
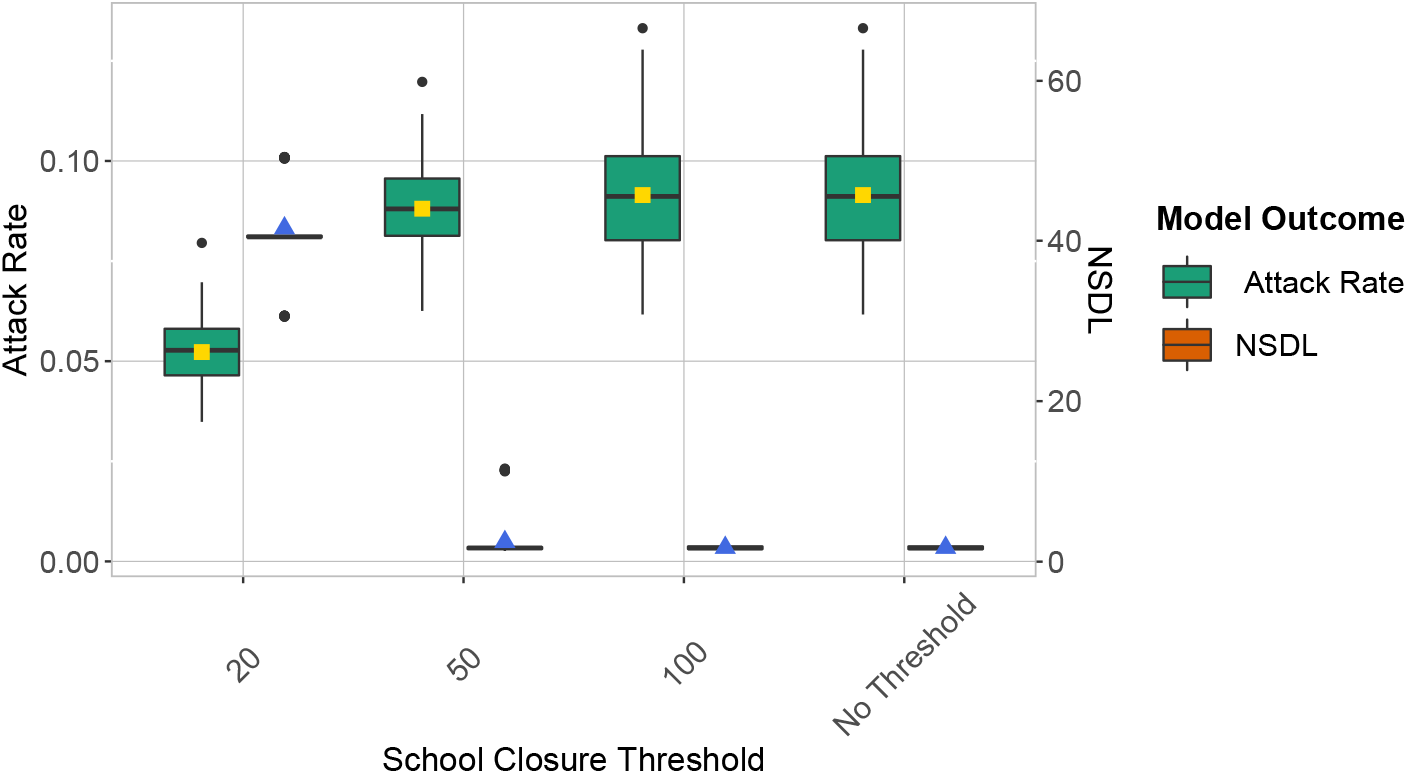
We show the repetitive testing strategy in the context of the Delta VoC for a moderate seeding of 5 seeds per week, where we consider different school closure thresholds. The class closure threshold is set to 8 detected cases. This experiments shows that a low school closure threshold has an effect on the attack rate (left panel) and it greatly reduces the NSDL (right panel).

## Footnotes

1 We refer to the Supplementary Information for a full description of the model.

2 https://www.sciensano.be/sites/default/files/report_seroprev_sars-cov-2_schools_t4_oct2021.pdf

3 Proportion of the infections generated in the school population, excluding seeded cases.

4 We assume a probability of asymptomatic infection of 0.8, therefore, we have the number of asymptomatic generations prior to encountering a symptomatic case by considering a geometric distribution’s mean, which is given by 1*/p*, where *p* = 1 − 0.8.

5 https://www.agodi.be/nieuwe-omkadering-basisonderwijs

6 https://www.vlaanderen.be/publicaties/vlaams-onderwijs-in-cijfers

## Notes

### Competing Interest Statement

The authors have declared no competing interest.

### Funding Statement

L.W and P.J.K.L. acknowledge support from the Research Foundation Flanders (fwo.be) (postdoctoral fellowships 1234620N and 1242021N). N.H. and P.J.K.L. acknowledge support from from the European Research Council (ERC) under the European Union Horizon 2020 research and innovation programme (grant number 101003688 EpiPose project). N.H. and A.T. received funding from the European Research Council (ERC) under the European Union Horizon 2020 research and innovation programme (grant number 682540 TransMID project). N.H. acknowledge funding from the Antwerp Study Centre for Infectious Diseases (ASCID) and the chair in evidence-based vaccinology at the Methusalem-Centre of Excellence consortium VAX-IDEA. C.M. received funding from the Fondation Leon Fredericq and the Fond de investissement de recherche scientifique from the CHU of Liege. G.D. received Postdoctorate Clinical Master Specialists funding from the Fund for Scientific Research (frs-fnrs.be). We used computational resources and services provided by the Flemish Supercomputer Centre (VSC) funded by the FWO and the Flemish Government. The funding agencies had no role in study design data collection and analysis decision to publish or preparation of the manuscript.

